# Identifying socio-economic barriers to antibiotic resistance stewardship in the Dairy Industry in LICs and LMICs

**DOI:** 10.1101/2025.01.20.25320874

**Authors:** Harshita Singh, Awanish Kumar Singh, Gargi Singh

**Affiliations:** Department of Civil Engineering, Indian Institute of Technology, Roorkee - 247667, India; College of Veterinary & Animal Science, G.B. Pant University of Agriculture and Technology, Pantnagar, Udham Singh Nagar – 263145

## Abstract

Antibiotic resistance (ABR) is a severe global public health threat, particularly in low- and middle-income countries (LICs and LMICs), where socio-economic and infrastructural factors impede effective ABR management. Here, we identify critical challenges, including disparity in the volume of research related to ABR stewardship for the dairy industry, that hinder the success of national action plans in LICs and LMICs. We conducted a literature review to pinpoint specific challenges to ABR stewardship in LICs and LMICs and employed generalised linear models to evaluate the predictive influence of dairy-related, socio-economic, and environmental factors on progress in antimicrobial resistance stewardship using TrACSS scores. We ascertained six key themes in otherwise interconnected challenges that critically limit ABR stewardship in LICs and LMICs: (i) lack of awareness among stakeholders, (ii) ineffective regulations, (iii) insufficient surveillance and monitoring, (iv) inadequate veterinary health infrastructure, (v) economic constraints, and (vi) certain cultural beliefs and traditional practices. Generalised linear regression models confirmed that effective surveillance, robust health and diagnostic infrastructure, and economic affluence significantly determine the progress of ABR stewardship in a country, as reported by TrACSS scores. However, the substantially sparse knowledge on the impact of dairy farms on environmental resistome generated in LICs and LMICs raises the concern that the interventions for the dairy industry in these countries are not informed by the actual local and national conditions.

## Introduction

Antimicrobial resistance (AMR) is a global public health threat, with estimates of approximately 4.95 million deaths associated with bacterial AMR in 2019.(Singer *et al*., 2016; Murray *et al*., 2022) The human-impacted environments, such as dairy farms, are the largest and ever-worsening reservoir of antimicrobial resistance, particularly to antibiotics.(Guo *et al*., 2021; Samreen *et al*., 2021) The misuse and overuse of antibiotics in the livestock industry are amongst the major contributors to the environmental resistome.(United Nations Environment Programme, no date; Holmes *et al*., 2016) Globally, it is estimated that 73% of all antibiotics are used in livestock rather than human healthcare, heightening concerns about antibiotic resistance (ABR) driven by practices in animal husbandry.(Van Boeckel *et al*., 2017; Marutescu *et al*., 2022) The dairy industry, which holds the largest share of global livestock units(Van Boeckel *et al*., 2015; FAO, 2020; Tiseo *et al*., 2020), is projected to be the fastest-growing livestock sector in this decade. This expansion of the dairy industry expects an increase in global milk production by 17%, primarily driven by low- and middle-income countries (LICs and LMICs) such as India and Pakistan. (*OECD-FAO Agricultural Outlook 2023-2032*, 2023) While the developed countries expect to have only a 6% increase in milk production along with a slight (0.61%) decrease in herd size, the developing countries expect to see a 19% increase in herd size and a concomitant 27% rise in milk production by 2032 (Table S1).(*OECD-FAO Agricultural Outlook 2023-2032*, 2023) The projected increase in herd size in developing countries is expected to increase the demand for antibiotics, further aggravating ABR-related challenges in LICs and LMICs, which are already grappling with high infectious disease burden.(Zhou *et al*., 2016; Huang *et al*., 2019; Guo *et al*., 2021; Zhao *et al*., 2021; Ardakani *et al*., 2023)

We know now that operational and infrastructural issues related to hygiene, sanitation, and biosecurity in dairy farms affect the disease prevalence and, thus, the antibiotic consumption and ABR patterns in LICs and LMICs.(Collignon *et al*., 2018; Ramay *et al*., 2020; Caudell *et al*., 2022; Singh *et al*., 2024, no date)However, there is sparse literature on the role of dairy farms in the environmental dissemination of antibiotic resistance from LICs and LMICs(Khan *et al*., 2020; Paul *et al*., 2021), leading to a reliance on projections from high-income (HICs) and upper-middle-income countries (UMICs), which may not accurately pinpoint the drivers of ABR in dairy farms in LICs and LMICs.(Van Boeckel *et al*., 2015; Mbaye *et al*., 2019; *Global Antimicrobial Resistance and Use Surveillance System (GLASS) Report 2022*, 2022; Sohaili, Asin and Thomas, 2024)

In HICs and UMICs, ABR stewardship in the dairy sector has been effective due to stringent regulations, advanced veterinary healthcare systems, and sophisticated farm management practices. However, the same has not been the case for LICs and LMICs, where higher disease prevalence, economic constraints, and inadequate veterinary infrastructure pose significant barriers to the successful implementation of strategies that have succeeded in wealthier nations.(Byarugaba, 2004; Ayukekbong, Ntemgwa and Atabe, 2017; Caudell *et al*., 2017; Adekanye *et al*., 2020; Cobo-Angel *et al*., 2021; Jani *et al*., 2021; Khan *et al*., 2022) The recommendations for ABR stewardship need to account for differences in dairy farming practices and infrastructure of LICs and LMICs. While wealthier countries can successfully focus on restricting unnecessary antibiotic use and raising awareness of ABR among farmers, LICs and LMICs must first tackle the higher prevalence of infectious diseases in farm animals and the under treatment of these animals due to socio-economic constraints.(Mbwasi *et al*., 2020; Pagani *et al*., 2020; Pattis *et al*., 2022; Portillo-Gonzalez *et al*., 2024) The stark contrast between the infrastructure and socio-economic conditions in which the dairy industry operates in HICs and UMICs, as opposed to LICs and LMICs, underscores the need for customised action plans for the latter.

We reviewed the literature to highlight the gap in research on the contribution of the dairy sector to the release of ABR in environment LICs and LMICs that limit the design of effective interventions. We then used a generalised linear model to identify key socio-economic, dairy industry- and climate-related factors significantly affecting ABR stewardship efforts in the veterinary sector. Finally, we identified several challenges unique to LICs and LMICs hindering National Action Plans (NAPs) based ABR stewardship in the veterinary sector, including lack of awareness, ineffective regulatory frameworks, insufficient surveillance and monitoring systems, inadequate access to veterinary healthcare, economic constraints, and traditional cultural practices, all of which limit the development of actionable guidelines within NAPs.

### Methodology

#### Literature review

To compare the volume of literature on environmental dissemination of ABR via dairy farms in LICs and LMICs vs. HICs and UMICs, we used two distinct queries (Query 1 and Query 2) on the Web of Science database (Clarivate Analytics) in May 2024. For Query 1, the following query was employed: “dairy AND (antimicrobial OR antibiotic resistance) AND f$cal OR dung OR manure OR wastewater NOT milk NOT (pig OR swine) NOT chicken”. This search returned 437 results, excluding the review articles. Each of these articles was individually screened to identify studies that either (i) used quantitative PCR to compare levels of antimicrobial resistance genes (ARGs) or (ii) reported culture-based results for antibiotic susceptibility analysis. Studies on microcosms or bench-scale experiments were excluded to ensure real-world relevance. Consequently, 30 articles reporting genetic data related to ABR and 50 articles detailing culture-based antibiotic susceptibility results were included in the assessment. For Query 2, the following search term: “dairy AND (antibiotic OR antimicrobial resistance) AND (infrastructure OR management OR operations) NOT (pig OR swine) NOT chicken”. This search returned 1002 results, excluding the review articles. Relevant articles were reviewed to identify challenges in the dairy industry in LICs and LMICs that may impede effective ABR stewardship. References within these articles were also examined for additional relevant literature.

#### Review of the National Action Plans (NAPs)

The NAPs of the top ten countries with the highest dairy production and largest herd were obtained from the WHO AMR library(WHO, no date) on May 23, 2024 (Table S2). The NAPs in non-English languages (Brazil, Russia, Argentina, and Chad) were machine-translated using Google Translate. The NAP for Mexico was unavailable in the WHO AMR library at the time of the search. HS manually reviewed the NAPs twice, focusing on antibiotic stewardship in the veterinary sector. Additionally, the country-wise most recent report for Tracking AMR Country Self-assessment Survey (TrACSS)(FAO *et al*., no date) was assessed to evaluate the status of the activities and legislations with a focus on the veterinary sector.

#### Statistical test

All the statistical tests were performed in R version 3.6.3(R Core Team, 2022) with RStudio(RStudio Team, 2023) 2023.03.0+386 “Cherry Blossom” Release for Windows as its graphical user interface. The significance of disparities in research output across countries from different income groups was assessed using a Chi-squared test for independence (p<0.05). The relative levels of antibiotic-resistant genes reported in the literature were tested for normality using the Shapiro-Wilk test. Since the data was not normal, the Kruskal Walis test (p<0.05) and the Wilcoxon rank sum test (p<0.05) were used.

Initially, a model with **TrACSS score** as the dependent variable and the dairy-related factors (**Milk** and **Head**); socio-economic factors (**Lit**, **GHSI** and **GDP**) and environmental factor (δ**T**) as the independent variables (Model 1). The assumptions of linearity, independence of errors, homoscedasticity, and normality of residuals were checked to ensure the validity of the model. The statistical significance of each predictor was assessed based on its corresponding t-value and p-value. The overall model fit was evaluated using the adjusted R^2^ and the F-statistic.

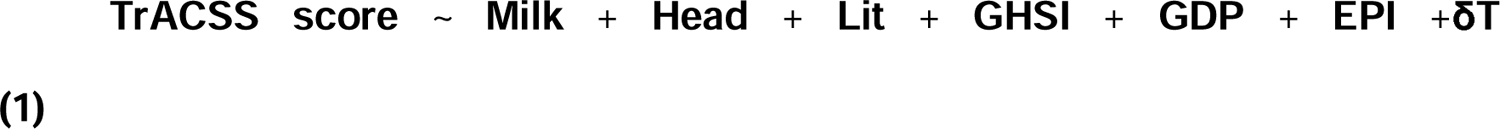

where,

#### TrACSS Score

The most recent country-wise Tripartite AMR Country Self-Assessment Survey (TrACSS) report was sourced from the Global Database.(FAO et al.) Only responses to questions addressing actions and legislation related to antibiotic consumption in the veterinary sector were used to calculate the TrACSS score (Figure 1B, 3C).

**Figure 1:**
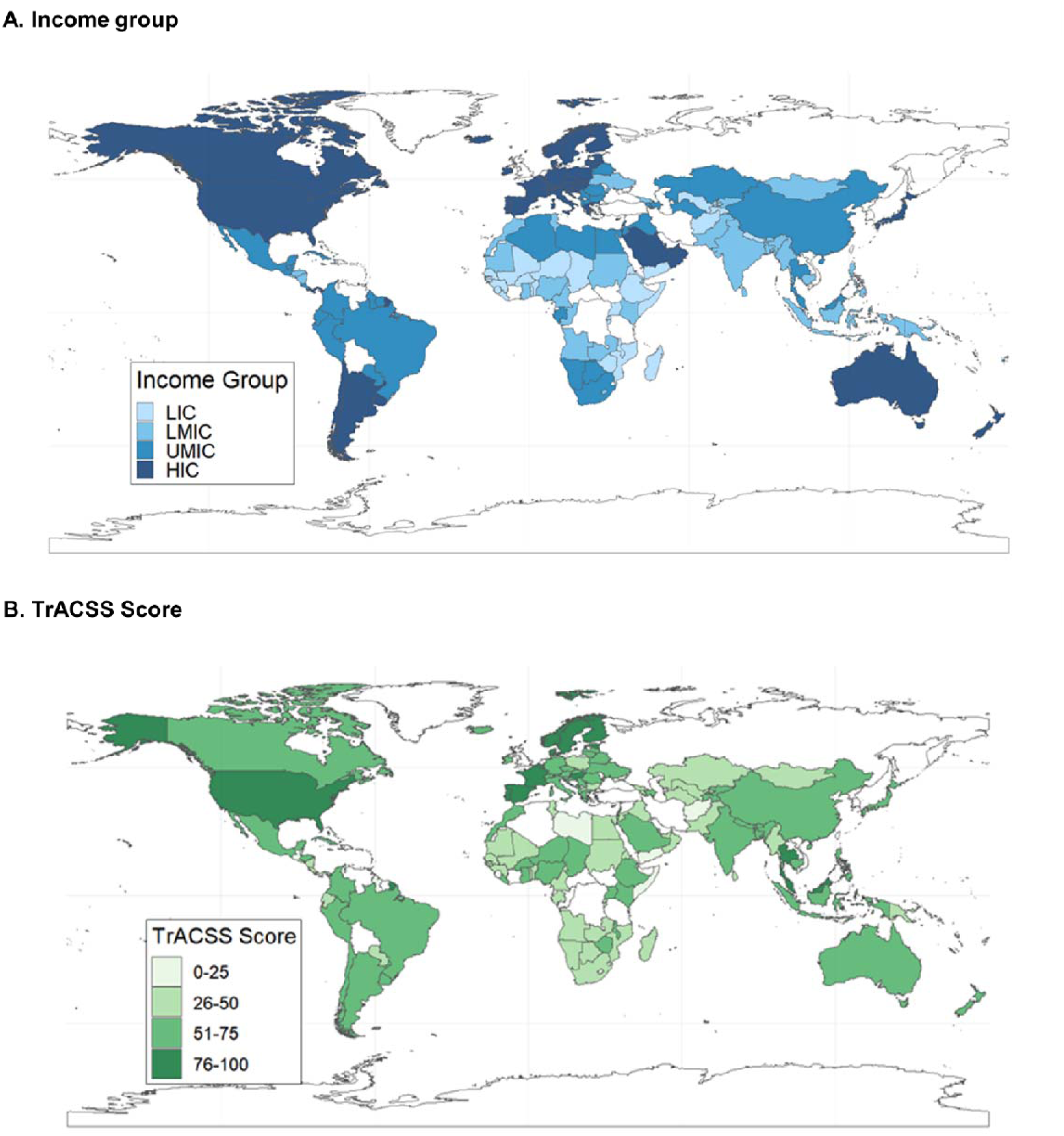
In sub-figure A, the map shows countries based on the income group defined by the World Bank. In sub-figure B, the map shows the TrACSS score of the countries based on their AMR stewardship in the veterinary sector.

**Milk:** Total country-wise cattle milk production (in million tonnes) for 2022.(FAOSTAT, 2022)

**Head:** Country-wise total herd inventory (in millions) for 2022.(FAOSTAT, no date a)

**Lit:** Literacy rate, based on the most recent available data(UNESCO Institute of Statistics (UIS), no date),

**GHSI**: Global Health Security Index, 2021(Bell and Nuzzo, 2021)

**GDP**: Gross domestic product per capita (USD)(FAOSTAT, no date b),

**EPI:** Environmental Performance Index 2024(Environmental Performance Index 2024 *With support from the McCall MacBain Foundation*, no date)

δ**T**: Change in surface temperature from 1961 to 2021(FAOSTAT, no date c)

After identifying GHSI as a significant predictor, the second model (Model 2) was developed to explore the categories constituting the fixed predictor variable:

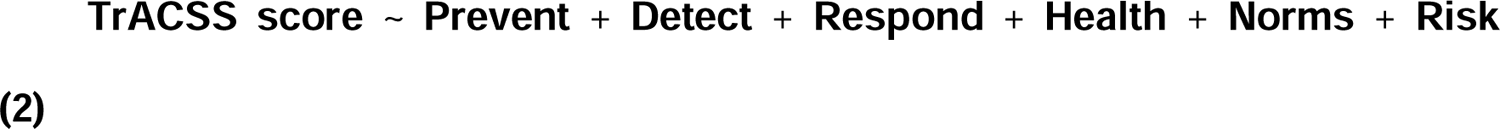

where,

**Prevent:** “Prevention of the emergence or release of pathogens”,

**Detect:** “Early detection and reporting epidemics of potential health concerns”,

**Respond:** “Rapid response to and mitigation of the spread of an epidemic”,

**Health:** “Sufficient and robust health sector to treat the sick and protect health workers”,

**Norms**: “Commitments to improving national capacity, financing and adherence to norms”, and

**Risk:** “Overall risk environment and country vulnerability to biological threats”.

Further, three separate models (Models 3, 4, and 5) were run to evaluate the association of indicators of the significant categories from the GHSI with the TrACSS score.

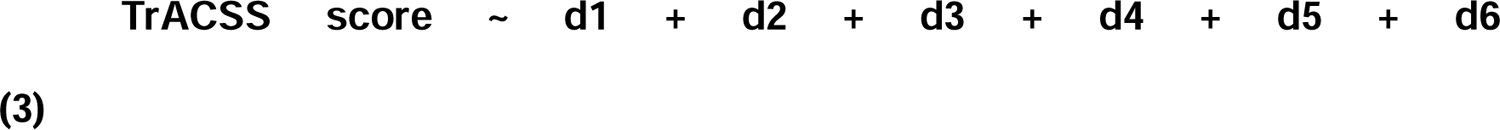

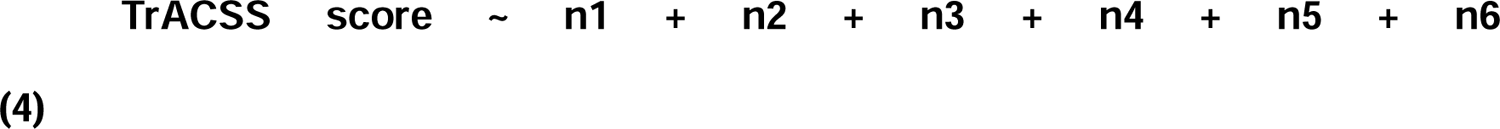

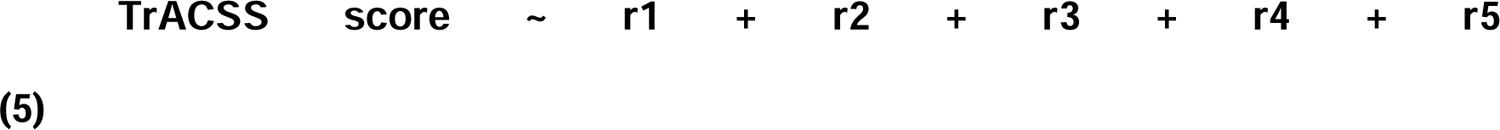

The details of the indicators in equations (3), (4), and (5) are detailed in SI Section S1. Model 3 focuses on factors related to laboratory systems, surveillance capabilities, data transparency, case investigations, and the epidemiology workforce. Model 4 examines international health commitments, including regulatory compliance, cross-border agreements, financing, performance of veterinary services, and data sharing. Model 5 addresses political risks, socio-economic resilience, infrastructure adequacy, environmental risks, and public health vulnerabilities.

## Results and Discussion

### Disparity in the volume of related research in LICS, LMICs, UMICs and HICs

For each query on Web of Science, the number of research papers were categorised into different groups of countries based on the economy (LIC, LMIC, UMIC, and HIC as defined by the World Bank) of the country (Figure 1A) that the author/s were affiliated.(World Bank Data, 2023) This allowed us to note the distribution of volume of research output across different economic contexts. For Query 1, 425 results were from HICs and UMICs, while only 48 were from LICs and LMICs (Figure 3D). Similarly, for Query 2, 999 results were from HICs and UMICs, while only 129 were from LICs and LMICs (Figure 3D). The sum of these figures is higher than the total number of results yielded per query (Query 1=437, Query 2=1002) because Web of Science filters results based on author affiliations and collaborative studies involving authors from multiple countries were counted once for each affiliated country leading to a duplication in counting.

**Figure 2:**
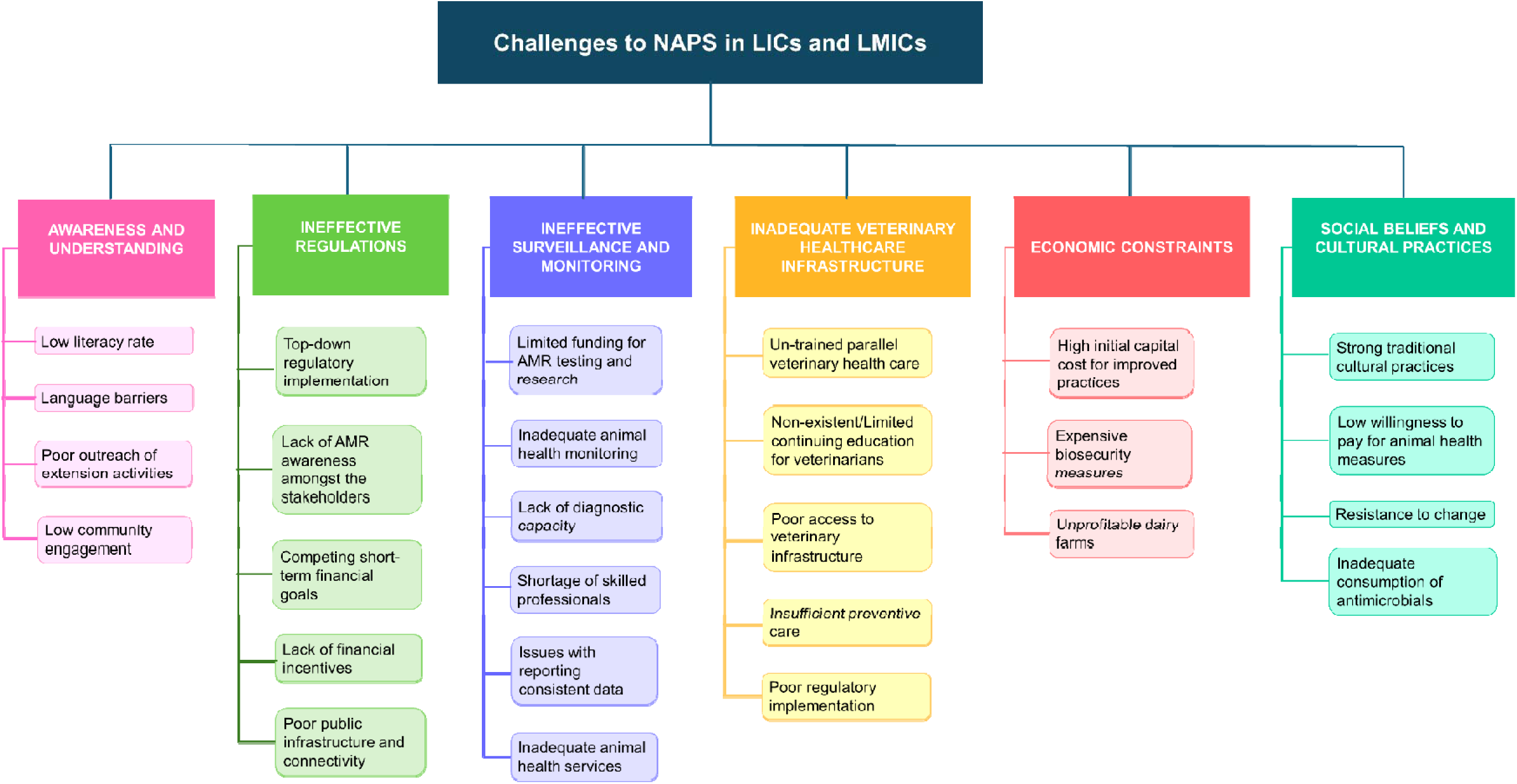
Challenges encountered in the veterinary sector, particularly within the dairy industry, regarding the implementation of National Action Plans (NAPs) for antimicrobial resistance (AMR) in Low-Income Countries (LICs) and Lower-Middle-Income Countries (LMICs).

**Figure 3:**
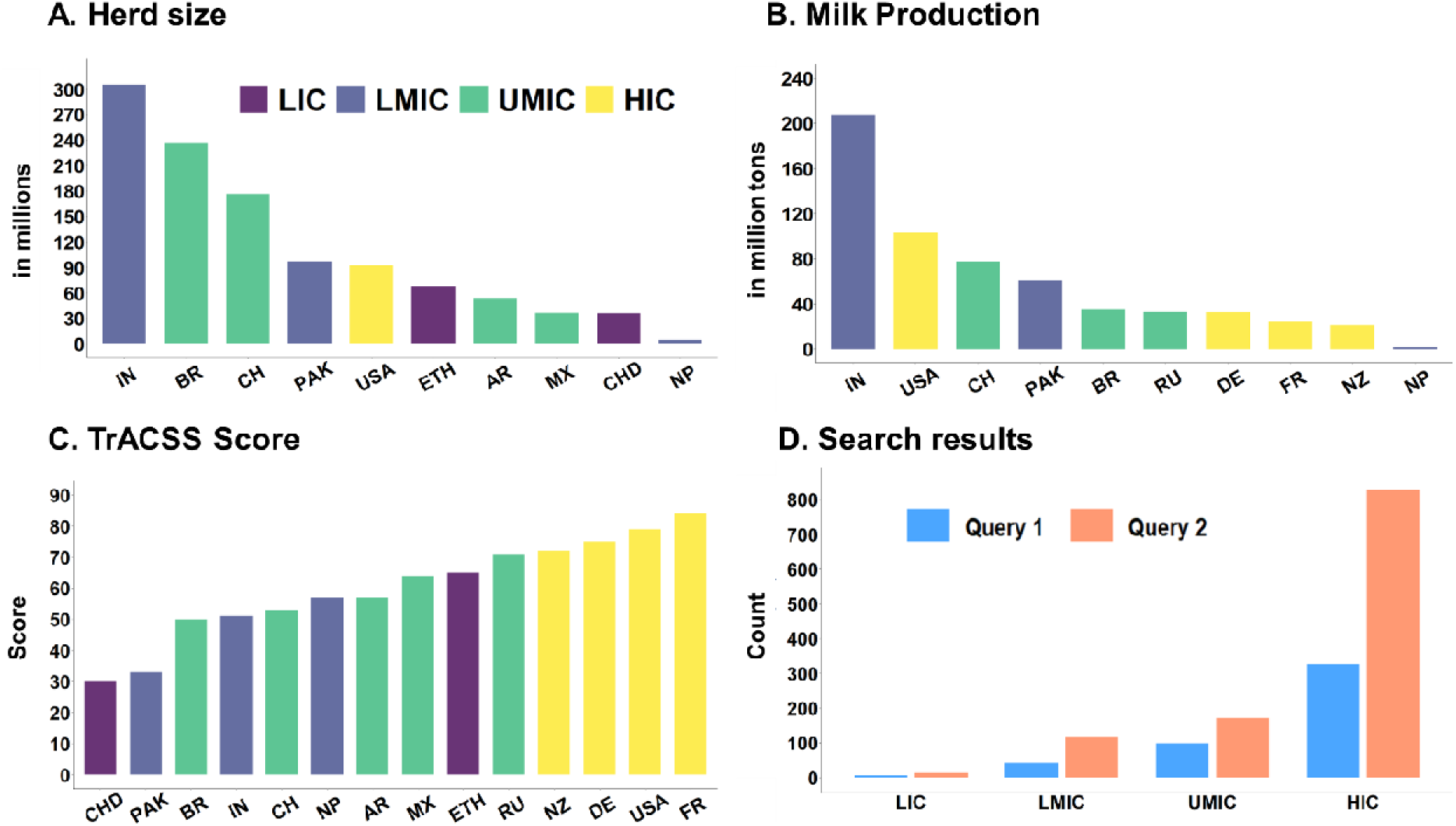
In sub-figures **A** and **B**, the y-axis represents herd size (in millions) and milk production (in million tons), respectively, across the top ten dairy-producing countries. The y-axis in sub-figure **C** displays the TrACSS score, derived from the assessment of country progress in AMR stewardship across the animal, food, agriculture, and legislation sectors for the year 2022. The x-axis in **A, B,** and **C** lists the countries: Chad (CHD), Pakistan (PAK), Brazil (BR), India (IN), China (CH), Nepal (NP), Argentina (AR), Mexico (MX), Ethiopia (ETH), Russia (RU), New Zealand (NZ), Germany (DE), the United States (USA), and France (FR). Sub-figure **D** presents the count of search results from the Web of Science database for each query, with the y-axis showing the result count and the x-axis categorising countries by income group as defined by the World Bank in 2022.

As reported elsewhere, we found a significant disparity in research output, with a much higher proportion of studies originating from wealthier nations (p-value=0.04, Chi-Square test).(Gozdzielewska *et al*., 2020) This disparity highlights a significant gap in data from lower-income regions, not because antibiotic resistance is absent in these nations but due to limited research capacity and reporting. The assumption that the policies that are successful in wealthier countries will be equally effective in LICs and LMICs overlooks the structural barriers in these regions and may inadvertently cause harm. As a result, transferring and applying the interventions and public policies designed for wealthier countries to economically weaker regions without first generating local knowledge risks unintended consequences due to distinct socio-economic and infrastructural features of LICs and LMICs. For instance, a study from Ethiopia reported widespread misconception among veterinary doctors that drug-resistant gram-positive infections, particularly MRSA, posing a greater threat than drug-resistant gram-negative infections, despite clear evidence of the high prevalence of highly resistant gram-negative pathogens.(Gebretekle *et al*., 2018) This misperception, likely influenced by familiarity with MRSA literature from other countries, appears to drive the inappropriate use of certain antibiotics. More broadly, this instance reveals a critical disconnect between clinical knowledge and actual practice in parts of the world, guidelines derived from high-income countries adversely affect prescribing behaviours in LICs and LMICs, ultimately undermining effective stewardship efforts.

Due to a lack of research output from LICs and LMICs, unique challenges for these nations have not been adequately addressed by policy makers. For example, public policies and interventions designed for HICs and UMICs are often supported by advanced healthcare and veterinary infrastructure, which is frequently absent in resource-limited settings of LICs and LMICs. This disparity can lead to policy bias, where researchers and policymakers overlook the ground realities of less affluent regions, resulting in misguided interventions that fail to address the root causes of ABR in these settings. For instance, in many LICs and LMICs, antibiotic misuse is driven more by the need to address visible disease symptoms in the absence of trained veterinary care rather than by want for growth promotion or even therapeutic attempts by trained care providers.(Khan *et al*., 2020; Frumence *et al*., 2021; Qijia Chua *et al*., 2021)

The regulatory crackdown on the self-determination and administration of treatment, as is frequently recommended, would curb any misuse or abuse of antibiotics and result in the loss of any chances of treatment for many sick animals. ABR stewardship strategies to be tailored to local contexts and invite active engagement from stakeholders, grounding the policies and interventions on local knowledge and data is indispensable.

### Environmental load of ABR from the dairy industry

Of the studies reporting the resistance of bacterial strains isolated from the samples of dairy excrements or their receiving environment (Query 1, n=50), 35 were from HICs, 9 from UMIC, 6 from LMICs, and 1 from an LIC (File S2, Figure 3B). Data on antibiotic resistance were available for *E. coli*, *Enterococcus spp*., *Salmonella spp.,* and *Campylobacter spp*., with only the data of *E. coli* being available from countries in each income group. The resistance patterns for *E. coli* (Figure 4A) showed that antibiotic resistance levels for different antibiotic classes, except for carbapenems (p-value = 0.025; HICs ∼ LICs), fluoroquinolones (p-value = 0.019; HICs ∼ LICs), and penicillin (p-value = 0.036; HICs ∼ LMICs), were comparable between developed countries (HICs and UMICs) and developing countries (LICs and LMICs). However, the percentage of *E. coli* isolates resistant to carbapenems, fluoroquinolones, and penicillin were higher in LMICs compared to HICs. A total of 31 studies reported the levels of antibiotic-resistant genes in dairy excrements and the receiving environments (surface/ground water and soil), with none originating from LMICs or LICs (Figure 4B). No significant difference was observed in the levels of the most abundantly reported antibiotic-resistant genes between the HICs and UMICs.

**Figure 4:**
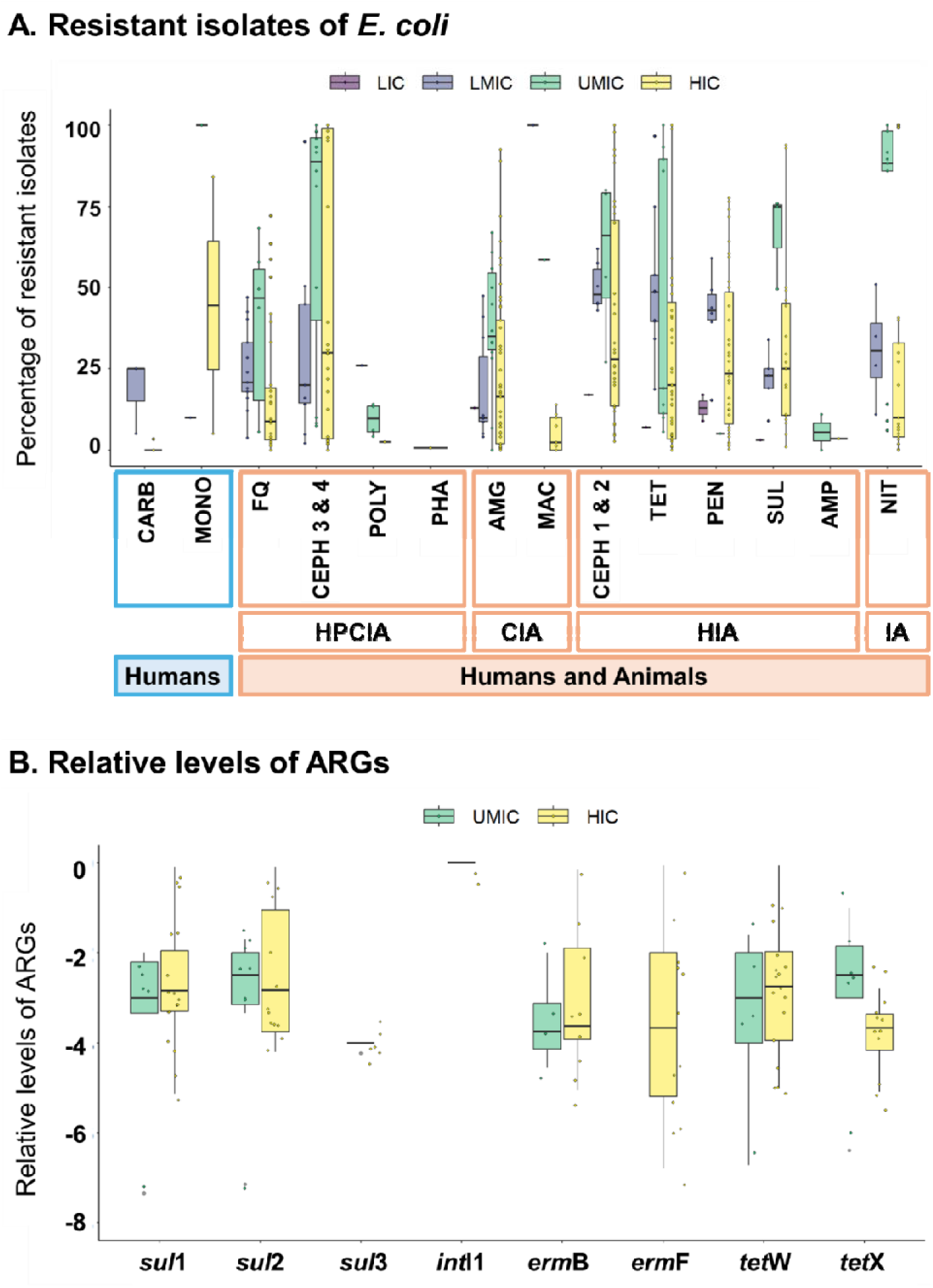
In sub-figure A, the y-axis represents the percentage of E. coli isolates resistant to various antibiotics reported from dairy excrement samples. The x-axis categorises these antibiotics into those recommended by the WHO for human use only ( CARB-Carbapenems, MONO-Monobactams) and those used for both humans and animals (FQ-Fluoroquinolones, CEPH 3&4-Cephalosporins 3^rd^ and 4^th^ generation, POLY-Polymyxins, PHA-Phosphonic acid derivatives, AMG-Aminoglycosides, MAC Mcrolides, CEPH 1&2-Cephalosporins 1^st^ and 2^nd^ generation, TET-Tetracyclines, PEN-Penicillins, SUL-Sulfonamides, AMP-Ampicillin, NIT-Nitrofuran derivatives). In sub-figure B, the y-axis shows the relative abundance of antimicrobial resistance genes (ARGs), while the x-axis lists the specific ARGs analysed. The legend indicates the income groups of the countries, as defined by the World Bank. In sub-figure B, the y-axis represents the relative levels of ARGs (from literature), and the x-axis represents the ARGs.

While some ARGs and antibiotic-resistant bacteria are globally prevalent, the higher resistance levels to certain critical antibiotics in *E. coli* LMICs indicate that these regions are uniquely and possibly critically challenged in the context of ABR. In the context of the commonly higher burden of infectious diseases, the reports of higher levels of ABR in isolates from LICs and LMICs underscore the urgent need to call for funding and supporting research and surveillance in LICs and LMICs to identify the local socio-economic and regulatory drivers of ABR in the dairy industry.

### Socio-economic, Dairy industry-, and climate-related Determinants of ABR stewardship

We evaluated the impact of various socio-economic, dairy- and climate-related predictors of the modified TrACSS scores as self-reported evaluations of AMR stewardship at the national level for 190 countries (Table 1,2; Figures S1, S2). A linear regression model (eq. 1) was developed with the TrACSS score as the dependent variable the following dairy-related factors as the independent variables-Total milk production (**Milk**) and Total herd inventory (**Head**) which are a measure of the scale of the dairy industry of the country; socio-economic factors-Literacy rate (**Lit**) which was used as an indicator for the awareness, **GHSI** is a measure of a country’s health security preparedness, and Gross domestic product per capita (**GDP**) reflecting the economic status of the country; and environmental factor-change in surface temperature (δ**T**) used as a proxy for global warming, which affects milk production and disease prevalence. The model demonstrated that the **GHSI** (slope = 1.195, p-value = 4.1e-11) was a significant predictor of the TrACSS score of a country (Table SX1, Fig X). Another model (eq. 2) evaluating the association of TrACSS score with the categories defining the GHSI was fit to identify the key factors within GHSI. Among these categories, **Detect** (slope=0.2281, p-value=0.03), **Norms** (slope = 0.2871, p-value = 0.01), and **Risk** (slope = 0.2227, p-value = 0.03) were positively associated with the TrACSS score (Table SX2, Fig X).

**Table 1:** Results for the linear regression models.

|  | **Model 1** | **Model 2** | **Model 3** | **Model 4** | **Model 5** |
| --- | --- | --- | --- | --- | --- |
| **Residual Standard error** | 17.42<br>(132 df) | 13.71<br>(164 df) | 14.21<br>(162 df) | 14.22<br>(163 df) | 15.74<br>(164 df) |
| **Multiple R-squared** | 0.3257 | 0.5204 | 0.4882 | 0.4875 | 0.3679 |
| **Adjusted R-squared** | 0.2899 | 0.5028 | 0.4692 | 0.4686 | 0.3486 |
| **F-statistic** | 9.106<br>(7,132 df) | 29.66<br>(6,164 df) | 25.75<br>(6,162 df) | 25.84<br>(6,163 df) | 19.09<br>(5,164 df) |
| **p-value** | 3.78E-09 | 2.20E-16 | 2.20E-16 | 2.20E-16 | 6.08E-15 |

**Table 2:** Significant variables for each linear regression model.

| Model 1: TrACSS score ~ Milk + Head + Lit + GHSI + GDP + EPI + cc |  |  |  |  |
| --- | --- | --- | --- | --- |
| Parameter | Estimate | Std. error | t-value | p-value |
| **GHSI** | 1.16E+00 | 1.80E-01 | 6.44 | 2.00E-09 |
| Model 2: TrACSS score ~ Prevent + Detect + Respond + Health + Norms + Risk |  |  |  |  |
| Parameter | Estimate | Std. error | t-value | p-value |
| **Detect** | 0.2281 | 0.1045 | 2.184 | 0.0304 |
| **Norms** | 0.2871 | 0.114 | 2.518 | 0.0128 |
| **Risk** | 0.2227 | 0.1035 | 2.152 | 0.0329 |
| Model 3: TrACSS score ~ d1 + d2 + d3 + d4 + d5 + d6 |  |  |  |  |
| Parameter | Estimate | Std. error | t-value | p-value |
| **d1** | 0.22611 | 0.04927 | 4.589 | 8.89E-06 |
| **d4** | 0.04534 | 0.03801 | 4.491 | 1.34E-05 |
| Model 4: TrACSS score ~ n1 + n2 + n3 + n4 + n5 + n6 |  |  |  |  |
| Parameter | Estimate | Std. error | t-value | p-value |
| **n2** | 0.09415 | 0.03142 | 2.997 | 0.00316 |
| **n3** | 0.35772 | 0.03768 | 9.494 | 2.00E-16 |
| Model 5: TrACSS score ~ r1 + r2 + r3 + r4 + r5 |  |  |  |  |
| Parameter | Estimate | Std. error | t-value | p-value |
| **r2** | 0.53471 | 0.11558 | 4.626 | 7.52E-06 |

Further analysis through additional GLM models focused on specific subcomponents within the **Detect, Norms,** and **Risk** categories, identifying several significant predictors of TrACSS scores. (Tables TX, SX3, SX4, SX5, Fig X). Within the **Detect** category (eq. 3), **Laboratory systems strength (d1)** (slope = 0.22611, p-value = 8.89e-06), and **Surveillance data accessibility and transparency (d4)** (slope = 0.22650, p-value = 1.34e-05) were significant factors, highlighting the critical function of well-equipped diagnostic laboratories in detecting resistant pathogens, facilitating timely interventions, and informing appropriate treatment strategies. Countries with robust surveillance systems are better positioned to track resistance patterns, share data across sectors, and respond proactively to emerging ABR threats, which tend to be higher-income nations.

Under the **Norms** category (eq. 4), **Cross-border agreements on public health and animal health emergency response (n2)** (slope = 0.09415, p-value = 0.00316) and **International commitments (n3)** (slope = 0.35772, p-value = < 2e-16) were the significant variables for TrACSS score. Cross-border agreements are essential for managing the spread of resistant pathogens across national boundaries, enabling collaboration in surveillance, data sharing, and coordinated interventions. International commitments, such as adherence to global AMR action plans, are particularly influential as they align national policies with global standards, strengthen legal frameworks, and promote the One Health approach, integrating human, animal, and environmental health strategies.

In the **Risk** Category (eq. 5), **Socio-economic resilience (r2)** (slope = 0.53471, p-value = 7.52e-16) was a key determinant of the modified TrACSS score. Nations with stronger socioeconomic resilience can invest in critical resources, such as veterinary services, education, disease prevention, and targeted therapy, which are vital for reducing antibiotic misuse in the dairy sector. Overall, our results indicate the need for an integrated approach that includes robust health systems, transparent surveillance, high-quality laboratory infrastructure, and international commitments to ensure effective ABR stewardship at the national level.

### Challenges to NAPs in the LICs and LMICs

The NAPs from the leading milk-producing countries and those with the largest herd sizes had five key themes: improving awareness and understanding of ABR among stakeholders, strengthening surveillance and monitoring of antibiotic use and ABR, reducing the incidence of infection by enhancing biosecurity measures on animal farms, optimising the use of antibiotics, and sustainable investment. As listed in Table S3, in LICs and LMICs, the limited resources, insufficient infrastructure, and weak regulatory enforcement create significant barriers to effective ABR stewardship, thereby jeopardising the achievement of the intended outcomes.

In the following section, we examine in detail the specific challenges for LICs and LMICs with respect to the current recommendations in their NAPs, which are interconnected but can be broadly classified into six major themes (Figure 2):

#### Lack of awareness amongst the stakeholders

Antibiotic stewardship is known to be directly proportional to the education level of the farmers.(Chan *et al*., 2012; Eltayb *et al*., 2012; Dhayal *et al*., 2023) A study from Tanzania underscored the role of education and literacy as relevant factors weighing in on the success of policies and interventions in the target population.(Roulette *et al*., 2017) Farmers with higher levels of education are more likely to be aware of ‘antibiotics’ and ‘antimicrobial resistance’ and are more likely to employ responsible practices.(Alhaji and Isola, 2018; Dhayal *et al*., 2023) Numerous studies highlight the lack of information amongst the stakeholders of the dairy industry, viz. farmers, local pharmacies, and parallel healthcare workers and professionals, regarding diseases, misuse of antibiotics and its role in the spread of ABR.(Alhaji and Isola, 2018; Obaidat *et al*., 2018; Gemeda *et al*., 2020; Sharma *et al*., 2020) Multiple studies across LICs and LMICs have documented that dairy farmers can recognise pictures of antibiotics as commonly used medicines, but they are usually unaware of what an antibiotic is, what comprises its misuse and abuse, and how that contributes to the increased ability of pathogens to resist antibiotic therapy.(Alhaji and Isola, 2018; Cambaco *et al*., 2020; Mutua *et al*., 2020; V. Kumar and Meena, 2021) Even amongst veterinary professionals, the general understanding of resistance is poor, often due to limited or non-existent continuing education.(Odetokun *et al*., 2019; Fasina *et al*., 2020; Wangmoi *et al*., 2021) The widespread lack of awareness of ABR undermines surveillance and disease prevention efforts and complicates the implementation of other antibiotic stewardship initiatives, thereby impeding effective control and containment of ABR.

In contrast, stakeholders in HICs and UMICs are better informed about the responsible use of antibiotics and ABR-related risks due to stronger regulatory frameworks, comprehensive awareness initiatives, and more robust veterinary health infrastructure. Enhancing antimicrobial stewardship on livestock farms requires tailored training programs that account for the various local language barriers, literacy levels, and prior experience of the targeted stakeholders.(Caudell *et al*., 2022) However, awareness campaigns without improving the healthcare infrastructure and economic resources would not be successful and effective in LICs and LMICs.(Caudell *et al*., 2022) In some instances, the ABR awareness campaigns in communities already struggling with inaccessible veterinary care have met with hostility, often resulting in continued misuse of antibiotics.(Mangesho *et al*., 2021)

#### Inadequate veterinary healthcare infrastructure

LICs and LMICs have a low ratio of veterinarian to livestock, which at times is 20X less than that in HICs and UMICs (Caudell *et al*., 2020, 2022), and hence the ability to provide the required veterinary care and antibiotic stewardship is compromised.(Haider *et al*., 2017; Alhaji and Isola, 2018; Chauhan *et al*., 2018; Mutua *et al*., 2020; Orubu *et al*., 2020; Frumence *et al*., 2021; Dankar, Hassan and Serhan, 2022)

The unstructured nature of the dairy industry in LICs and LMICs presents logistical difficulties, such as the inability of veterinarians to visit farms or the impracticality of transporting dairy animals to healthcare centres. In many cases(Roess *et al*., 2015; Kumar and Gupta, 2018; Mutua *et al*., 2020), it is more affordable and convenient for dairy farmers to turn to pharmacies and parallel healthcare for medical attention.(Espadamala *et al*., 2018; Vikash Kumar and Meena, 2021; Garzon *et al*., 2023)

Informal treatment practices often address the therapeutic needs but can also result in failed treatment, contributing to the misuse of antibiotics or abuse of antibiotics through the administration of inappropriate or inadequate doses.(Sudhinaraset *et al*., 2013; Sharma *et al*., 2020) The unregulated dispensing of antibiotics without adequate guidance further complicates efforts to manage and mitigate ABR.(Barker *et al*., no date; Cheema *et al*., 2001; Kotwani *et al*., 2012) While this alternative system, driven by insufficient training and awareness, can unintentionally contribute to the rise of ABR, proper support and training could transform it into a valuable resource, bridging gaps in veterinary care and aiding efforts to combat resistance.(Caudell *et al*., 2020)

In the absence of parallel veterinary healthcare, farmers often resort to self-prescribing treatments for their animals, relying on personal experience and facilitated by the unrestricted availability of antibiotics without veterinary prescription and direct marketing of antibiotics to dairy farmers, a practice that poses a significant risk factor for the development of antibiotic resistance in the dairy sector.(Awad *et al*., 2005; Eltayb *et al*., 2012; Carrique-Mas and Rushton, 2017; Alhaji and Isola, 2018; Obaidat *et al*., 2018; Vijay *et al*., 2021; Vikash Kumar and Meena, 2021) This misuse is exacerbated by the misperception that certain pharmaceutical products contain sub-optimal levels of active ingredients, leading farmers to administer higher-than-recommended doses.(Alhaji and Isola, 2018) In their pursuit of prompt and observable improvements in livestock health, the farmers pressurise veterinarians to prescribe antibiotics, even in cases where such treatment may be unnecessary.(Chauhan *et al*., 2018; Mutua *et al*., 2020) This dynamic is fuelled by economic pressures faced by both farmers and veterinarians to maintain productivity and profitability. Given the economic risks for dairy farmers that a sick animal poses, any improvement in veterinary infrastructure and accessibility of veterinary care would have a positive impact not only on animal health and productivity but will also be a key step towards antibiotic stewardship in LICs and LMICs.

#### Ineffective regulations

NAPs aim to develop systems and adopt regulations to track antibiotic usage, monitor resistance patterns, and provide critical data for informing policy decisions and early detection of emerging resistance trends. In HICs and UMICs, the ABR stewardship is significantly strengthened by robust regulatory frameworks and well-developed veterinary infrastructure. Regulation, when access to veterinary treatment is available, can be very effective for controlling the misuse of antibiotics. For instance, in the Netherlands, stringent control and monitoring of antibiotic use in animals led to a 70% reduction in veterinary antibiotic sales and a marked decline in ABR associated with livestock.(NethMap 2022: Consumption of antimicrobial agents and antimicrobial resistance among medically important bacteria in the Netherlands, 2022)

However, in LICs and LMICs, the top-down regulatory approach to antibiotic stewardship has been largely ineffective. This failure is driven by high demand for over-the-counter antibiotics for therapy, often due to limited access to veterinary care, and compounded by weak regulatory enforcement caused by knowledge gaps in the design of policy and resource constraints.(Srivastava, 2011; DAHDF, 2013; MoHFW, 2016; Mutua *et al*., 2020; Casey *et al*., 2023) Any regulatory imposition on the over-the-counter sale of antibiotics must strike a balance between permitting therapeutic attempts in countries with high disease burden and low access to healthcare resources and curtailing the irresponsible use of antibiotics simultaneously. Without improvement in both the quality of medical and veterinary care and ease of access to them, the policy of regulating over-the-counter sale of antibiotics is bound to be ineffective. Instead, the measures to restrict the sale of antibiotics might worsen the widespread undertreatment and non-compliance, resulting in the loss of productive animals.(Ojo *et al*., 2016) On a larger scale, regulations such as a blanket ban on non-therapeutic antibiotic use, which have been effective in many developed countries, could have adverse repercussions for dairy farmers in developing nations, where the veterinary infrastructure required to discern therapeutic use from non-therapeutic use is inaccessible.(Ojo *et al*., 2016; Kakkar *et al*., 2018; Mishal S Khan, Sonia Rego and Julia Spencer, 2018)

Another challenge to the regulatory control on the sale and use of antibiotics comes from a lack of resources to ensure compliance, a recurring theme for all LICs and LMICs.(Chokshi *et al*., 2019) However, the regulatory challenges extend beyond to include the push for sale by the pharmaceutical companies via direct marketing and sale of antibiotics and even active pharmaceutical ingredients to farmers and small-scale feed manufacturers.(L Kumaranayake *et al*., 2000; Suleman *et al*., 2016; Sommanustweechai *et al*., 2018; Khan *et al*., 2020)

#### Insufficient surveillance and monitoring

Until 2020, the national-level surveillance systems monitoring antibiotic resistance in animals existed only in HICs and UMICs.(Diallo *et al*., 2020) These countries maintain comprehensive databases that track animal health at both the herd and national levels. Regular data on disease prevalence and the distribution and sale of antibiotics are collected from pharmacies, veterinarians, local distributors, and the pharmaceutical industry, which reflect on the scale of antibiotic usage and inform policy design.(Merle *et al*., 2014; Tree *et al*., 2024) However, the successful development and maintenance of comprehensive data collecting and recording systems require substantial financial investment and logistical support, which for many LICs and LMICs(Chauhan *et al*., 2016; Obaidat *et al*., 2018; Hussein *et al*., 2023), is a substantial barrier.(Khan *et al*., 2020; Iskandar *et al*., 2021) Monitoring antibiotic use in the dairy industry of LICs and LMICs is minimal.(L Kumaranayake *et al*., 2000; Sommanustweechai *et al*., 2018). The decentralised small-scale nature of dairy farming further complicates enforcement efforts(Obaidat *et al*., 2018; Qijia Chua *et al*., 2021), as monitoring numerous small-scale farms becomes logistically challenging and resource intensive.(Mishal S Khan, Sonia Rego and Julia Spencer, 2018; Cambaco *et al*., 2020) The inadequate monitoring and tracking of pharmaceutical ingredients and products, coupled with insufficient inspections at drug distributors and retailers, further complicate field-level surveillance and regulation.(Sommanustweechai *et al*., 2018)

In the dairy sector, antibiotics are often used without veterinary prescriptions, and even when sought with a prescription, no testing is done for pathogens or susceptibility analysis is seldom, leading to a lack of data on disease prevalence and resistance.(Ayukekbong, Ntemgwa and Atabe, 2017; Caudell *et al*., 2020) Veterinarians, concerned about farmers’ adherence to complex or extended regimens, often prescribe broad-spectrum antibiotics. While these antibiotics are effective against multiple infections, their use without adequate diagnostics can contribute to the development of antimicrobial resistance.(Kemp *et al*., 2021)

In countries with inadequate veterinarian-to-animal ratios, meeting the demand for diagnostic services and qualified professionals remains a significant challenge.(Fitzgibbon and Wallis, 2013; Ombelet *et al*., 2018; Frumence *et al*., 2021; Qiu *et al*., 2024) This leads to difficulties in designing effective and relevant policies in the absence of reliable and consistent data on disease prevalence and antibiotic susceptibility.

#### Economic constraints

The implementation of NAP interventions, such as enhanced surveillance and monitoring of ABR and antibiotic usage, alongside improved biosecurity measures, requires substantial economic investment.(Dik and Sinha, 2017; Samreen *et al*., 2021) A country’s “**socio-economic resilience**” significantly influences its self-reported success in veterinary ABR stewardship (model 4). Strengthening economic resilience and boosting available resources for capacity-building initiatives and biosecurity enhancements are crucial for effective ABR stewardship. Wealthier nations are generally better equipped to invest in new technologies and practices for effective ABR management.(Zhou *et al*., 2022; Farhan *et al*., 2024) In contrast, financially constrained countries face challenges in implementing operational changes to improve biosecurity on dairy farms, such as restricting the movements of sick animals, enforcing quarantine disinfection protocols and developing necessary infrastructure like diagnostic facilities.(Postma *et al*., 2016; Dhaka *et al*., 2023a) Limited resources restrict the ability to access professional veterinary care(Obaidat *et al*., 2018) even when these services are locally available.(Chauhan *et al*., 2016) In addition to the international differences in the availability of financial resources, the inequality within a country also impacts the ability to effectively implement interventions for all social and economic classes.

Thus, the ability to adopt biosecurity measures and access to veterinary care is further skewed in favour of the minority large-scale dairy farms, even when the country secures investment and prioritises antibiotic stewardship. The economic resources are unequally distributed and typically vary according to farm size (Dhaka *et al*., 2023b; Muloi *et al*., 2023), amongst other socio-economic factors.

In contrast to the concentrated animal feeding operations in HICs and UMICs(Beattie *et al*., 2018; Zhang *et al*., 2021; Usepa *et al*., 2022), in LICs and LMICs, much of the industry relies on small-scale dairy farms with as small as a single animal per farm. For small-scale farmers, the dairy animals often provide supplemental income and ensure nutritional security for their families.(Balehegn *et al*., 2020; Paul *et al*., 2021; Sekaran *et al*., 2021) Consequently, they rely on untrained parallel healthcare workers and only seek veterinary care when the animal’s condition becomes severe, usually after multiple antibiotics have been administered by untrained practitioners.(Kumar Mohanta, 2012; Chauhan *et al*., 2016; Sharma *et al*., 2020) Large-scale dairy farmers, on the other hand, tend to engage qualified veterinarians and use antibiotics appropriately.(Kumar and Gupta, 2018)Large-scale dairy farms are typically more likely to afford advanced waste management systems like flushed or vegetative treatment systems.(Durso, Miller and Henry, 2018; Wallace *et al*., 2018; Fan *et al*., 2020; Wang *et al*., 2021, 2022; Kempf *et al*., 2022)

Investment in advanced waste management technologies is almost always unfeasible for small-scale farmers who need to prioritise cost-effective solutions that align with their financial constraints and operational scale.(Katada *et al*., 2021) Biosecurity measures commonly implemented in large farms, such as footbaths, slated floors, and robotic cleaners, are also typically neither practical nor feasible for smaller farms. Consequently, smaller farms may struggle to achieve the same level of biosecurity, potentially leading to higher risks of pathogen transmission and, subsequently, a higher need for antibiotics.(Alam *et al*., 2019; Ikhimiukor *et al*., 2022) Limited financial resources pose a significant barrier to investing in sustainable farming practices, vaccines, and alternative treatments that could lessen reliance on antibiotics. At the individual level, these economic constraints frequently outweigh the efforts to improve awareness, optimise antibiotic use, and implement effective ABR mitigation strategies. Without robust economic justification and investment in these strategies, progress towards achieving sustainable antibiotic use, as envisioned by GAP-AMR, is severely hampered. At the same time, when the ABR stewardship-related recommendations are also aligned with economic incentives in LICs and LMICs, they are willingly followed. For example, it is more profitable for farmers to withhold the sale and consumption of milk post-antibiotic therapy(Attaie *et al*., 2015) when the antibiotics and their derivatives in milk interfere with the downstream fermentation process.(Obaidat *et al*., 2018) Discarding milk contaminated with antibiotic residues means foregoing essential income needed to support families. Many small-scale farmers also face challenges in isolating sick animals due to space and facility constraints, often resulting in the spread of infection and increased animal mortality rates.(Mutua *et al*., 2020)

#### Social beliefs and cultural practices

For small-scale farmers in LICs and LMICs, ethnoveterinary medicine plays an important role in animal production and livelihood development and is frequently the only option for farmers to treat their sick animals.(Iqbal *et al*., 2014; Aziz *et al*., 2018; Jayakumar *et al*., 2018a; Chaachouay *et al*., 2022; Rehman *et al*., 2022) In some regions, the cultural norms accord high status to the care of dairy animals, especially cows, making their care a socially rewarding act, which in resource-constrained societies with inaccessible veterinary care is a primary driver for the care of sick animals.(Ahmadi *et al*., no date). Similarly, some cultural beliefs of the farmers about the diseases could be a barrier to seeking treatment and effective health services. For instance, some pastoralists of Borana in Ethiopia believe that ‘*buda*’ (evil eye) was the cause of mastitis in their cattle which could be treated by *tufaa* (spitting).(Amenu *et al*., 2017a) On the other hand, the Masaai pastoralists of Tanzania have been documented to use antimicrobials as ‘energisers’ and ‘fatteners’ for the livestock.(Caudell *et al*., 2022)

Further, some operational practices in LICs and LMICs that are influenced by the local culture also increase the exposure of the dairy farm workers and the local community to the dairy-waste microbiome. For example, manually handling dung, which includes the production and use of cow dung cakes, and living in proximity of the dairy animals puts the local community at an elevated risk of direct exposure to the microbiome associated with dairy farms. In developing countries where an integrated crop-livestock system(Sekaran *et al*., 2021) is widespread, dairy farmers have frequent and close contact with food animals in line with local practices and social beliefs.(Okeke *et al*., 2005) Limited mechanisation of feeding, cleaning, and waste management further exacerbates the exposure to antibiotic resistance in the dairy farms of LICs and LMICs.(Lindahl *et al*., 2018) Additionally, small-scale subsistence farmers are more likely to rely on dairy products from their farms for nutrition and are more likely to be exposed to any antibiotic-resistant food-borne pathogens via ingestion.(Eltayb *et al*., 2012) Due to inadequate dairy waste management, which is common in LICs and LMICs, the dairy-related microbiome circulates in the local environment. With larger exposure to dairy products, it is not surprising that in LICs and LMICs(Katakweba *et al*., 2015; Afema *et al*., 2016), genotypic similarities have been documented between the resistant enteric bacteria in livestock and the nearby communities. On the other hand, in HICs and UMICs(Mather *et al*., 2012; Dorado-García *et al*., 2018; Day *et al*., 2019), they were distinguishable, and any overlap was dependent on the intensity of contact between the animals and the human population.

Given the existing public health burden from infectious diseases in these regions(Mbwasi *et al*., 2020), the increased exposure to any zoonotic pathogens in the dairy farms could exacerbate the community transmission of ABR, underscoring the need for better management practices and diagnostic facilities.

## Conclusion

ABR stewardship in LICs and LMICs is often challenged by limited awareness, inadequate surveillance, weak regulations, insufficient veterinary infrastructure, economic constraints, and prevailing beliefs. Addressing these challenges requires a comprehensive approach to enhance ABR management. The success of ABR-related policies requires feedback and information from farm managers and their veterinary consultants.(Afema, Davis and Sischo, 2019) This is more important in LICs and LMICs, where the interventions need to be cognizant of and informed by the socio-economic conditions at the level of the typical dairy farmer and involve all the stakeholders.(Khan *et al*., 2020; Singh *et al*., 2021) Rather than a top-down approach, a bottom-up approach would be more effective in the case of LMICs and LICs, similar to the social construction framework proposed by Legido-Quigley et al.(Legido-Quigley *et al*., 2019)

Fostering community engagement and awareness through organising community-based programs that involve farmers, local leaders, and other stakeholders in discussions about ABR and the importance of responsible antibiotic use is important. Regular focused group discussions on the use of local and social media and social networks to disseminate information and success stories on proper antibiotic use are needed to increase awareness of ABR and the responsible use of antibiotics. Translating behavioural intentions into sustained behaviour change would require continuous veterinary support, education, and incentives.(Jones *et al*., 2015; Amenu *et al*., 2017b; Farrell *et al*., 2023; Regan *et al*., 2023)

In regions with limited and struggling veterinary healthcare systems, training parallel healthcare workers to recognise common diseases in the dairy industry, monitoring antibiotic use, and knowing when to refer animals to trained veterinary professionals would be advantageous. This includes continuous education programs for veterinarians, para-veterinarians, and pharmacists on the recent ABR trends, diagnostic techniques, and treatment protocols. Certification programs that ensure veterinarians are proficient in ABR management and antibiotic stewardship need to be introduced. Such training of parallel healthcare providers for human health has demonstrated efficacy in improving clinical outcomes and reducing irrational antibiotic use.(Das *et al*., 2016)Also, ethno-veterinary medicine culture as an alternative must be explored and promoted to reduce the complete reliance on antibiotics for treatment.(Jayakumar *et al*., 2018b; Mutua *et al*., 2020)

The policies must also consider economic feasibility. For instance, implementing high-cost biosecurity measures like automated cleaning systems or advanced waste management technologies in smaller farms with limited resources may be impractical. Instead, low-cost and scalable solutions, such as regular handwashing, basic sanitation practices, and low-cost disinfection methods, could be more feasible and equally effective. Implementing funding mechanisms, including subsidies, to promote sustainable agricultural practices and foster economic resilience is crucial for tackling the intertwined issues of economically constrained dairy farming in LICs and LMICs. Community-based approaches, such as establishing centralised manure composting facilities, collective wastewater treatment infrastructure, and farmer field schools for training in sustainable practices, can collectively enhance biosecurity and hygiene standards while reducing antimicrobial resistance risks within a region.

An effective way to reduce the use of antimicrobials and thus prevent ABR is to decrease the need for these treatments in the first place, aka disease prevalence.(World Health Organization., 2012) Interventions to improve overall animal health and reduce disease prevalence by identifying and promoting best-practices and non-pharmaceutical interventions, such as maintaining good hygiene, providing proper nutrition, and ensuring optimal living conditions, are crucial in reducing their susceptibility to infections.(Pinto Ferreira *et al*., 2022; Singh *et al*., 2024) By focusing on these preventive measures, we can minimise the need for antimicrobial treatments, thereby helping to curb the rise of ABR.

In conclusion, a multifaceted and inclusive approach that incorporates socio-economic realities, stakeholder engagement, and practical interventions is essential to effectively address antimicrobial resistance in LMICs, ensuring the health and well-being of both animals and humans.

## Data Availability

All data produced in the present work are contained in the manuscript

## Acknowledgements

This research was supported by Anusandhan National Research Foundation (formerly SERB), India (Grant number: CRG/2020/005658) and Faculty Initiation Grant provided by IIT Roorkee (FIG/1007621).

## Authors contribution

Conceptualisation: Gargi Singh and Harshita Singh; Methodology: Gargi Singh; Formal analysis and investigation: Harshita Singh; Writing-original draft preparation: Harshita Singh; Writing-review and editing: Gargi Singh; Funding acquisition: Gargi Singh and Awanish Kumar Singh; Resources: Gargi Singh and Awanish Kumar Singh; Supervision: Gargi Singh and Awanish Kumar Singh

