## Supplemental Information 1 for "Identifying socio-economic barriers to antibiotic resistance stewardship in the Dairy Industry in LICs and LMICs"

**S1: Details of parameters used in Model 3, Model 4 and Model 5**

**Model 3: TrACSS score ~ d1 + d2 + d3 + d4 + d5 + d6**

**d1:** Laboratory systems strength and quality which includes Laboratory capacity for detecting priority diseases and Laboratory quality systems,

**d2:** Laboratory supply chains which includes Specimen referral and transport system and Laboratory cooperation and coordination,

**d3:** Real-time surveillance and reporting which includes Indicator and event-based surveillance and reporting systems and interoperable, interconnected, electronic real-time reporting systems,

**d4:** Surveillance data accessibility and transparency which includes Coverage and use of electronic health records, Data integration between human, animal, and environmental health sectors, Transparency of surveillance data, Ethical considerations during surveillance, and international data sharing,

**d5:** Case-based investigation which includes Case investigation and contact tracing and point of entry management,

**d6:** Epidemiology workforce which includes Applied epidemiology training program and Epidemiology workforce capacity.

**Model 4: TrACSS score ~ n1 + n2 + n3 + n4 + n5 + n6**

**n1:** International Health Regulations (IHR) reporting compliance and disaster risk reduction which includes Official IHR reporting and Integration of health into disaster risk reduction,

**n2:** Cross-border agreements on public and animal health emergency response which includes Cross-border agreements,

**n3:** International commitments which includes Participation in international agreements and Voluntary memberships,

**n4:** Joint External Evaluation (JEE) and Performance of Veterinary Services (PVS) Pathway which includes Completion and publication of a JEE assessment and gap analysis and Completion and publication of a PVS assessment and gap analysis,

**n5:** Financing which includes National financing for epidemic preparedness, Financing under Joint External Evaluation (JEE) and Performance of Veterinary Services (PVS) reports and gap analyses, Financing for emergency response, and Accountability for commitments made at the international stage for addressing epidemic threats,

**n6:** Commitment to sharing of genetic and biological data and specimens which includes Commitment to sharing genetic data, clinical specimens, and/or isolated specimens (biological materials) in both emergency and nonemergency research

**Model 5: TrACSS score ~ r1 + r2 + r3 + r4 + r5**

**r1:** Political and security risk which includes Government effectiveness, Orderly transfers of power, Risk of social unrest, Illicit activities by non-state actors, Armed conflict, Government territorial control, and international tensions,

**r2:** Socio-economic resilience which includes Literacy, Gender equality, social inclusion, public confidence in government, Local media and reporting, and Inequality,

**r3:** Infrastructure adequacy which includes adequacy of road works, airports, and power network,

**r4:** Environmental risks which includes Urbanization, Land use and Natural disaster risk,

**r5:** Public health vulnerabilities which includes access to quality healthcare, potable water and sanitation, Public healthcare spending levels per capita, and trust in medical and health advice.

**Table S1:** Net growth in milk production (kilotons per week) and herd inventory (herd size) globally, as well as for developed and developing nations individually, over the period from 2022 to 2032.

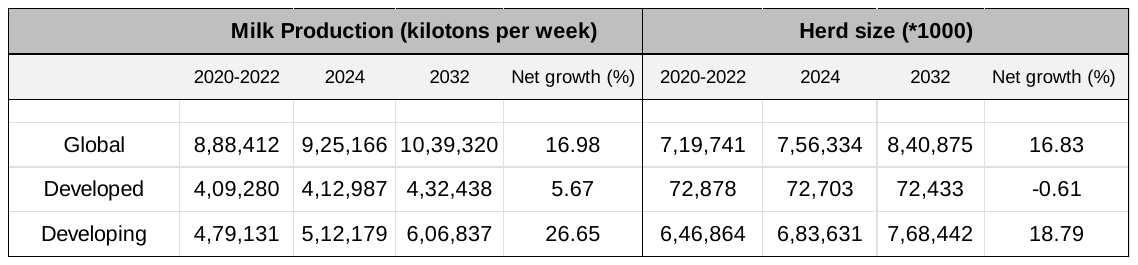

**Table S2:** Countries with the highest milk production and largest herd size in 2022 (FAOSTAT data).

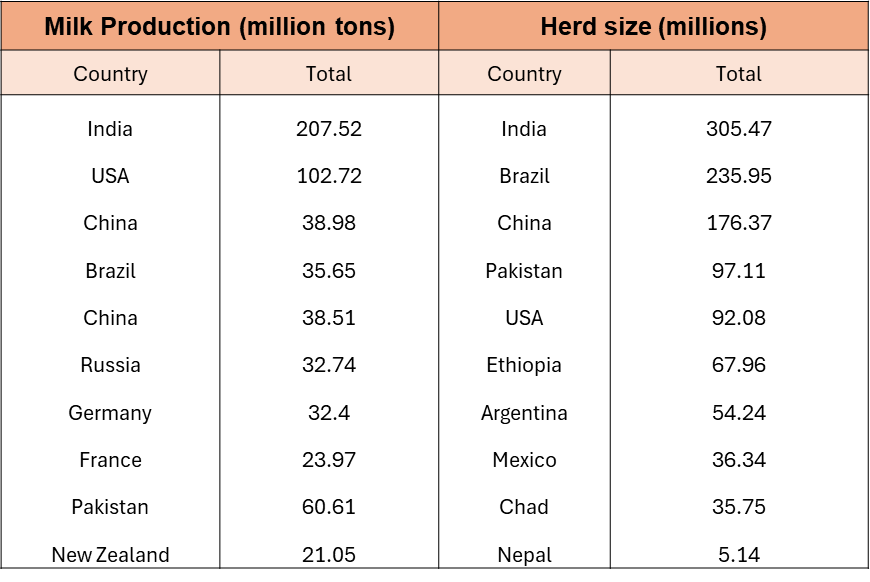

**Table S3:** Recommendations and Challenges for successful implementation of National Action Plans (NAPs)

| Recommendations | Challenges |
| --- | --- |
| Integration of Antimicrobial Resistance (AMR) topics into the curricula of veterinary education. This includes revising existing courses, developing new modules, and providing ongoing professional development opportunities. | Developing and distributing new educational materials and training programs require significant investment in time, money, and expertise. |
| Establishment or enhancement of surveillance systems to monitor AMR trends in veterinary health, agriculture, and the environment. This includes setting up laboratories, databases, and data collection frameworks. | Scattered, small holdings often make it difficult to monitor and survey. Many regions lack the necessary technological infrastructure and trained personnel to implement effective surveillance systems. Establishing and maintaining surveillance systems is costly, requiring sustained financial investment, which may be challenging in low-resource settings. |
| Implementation of IPC measures in the veterinary practices and food production to prevent the spread of infections and reduce the need for antibiotics. This includes education on hygiene practices, development of protocols, and monitoring adherence to standards. | Economic constraints would hinder the adoption and implementation of IPC. Lack of veterinary access would result in non-compliance. Ensuring consistent adherence to IPC protocols is challenging, especially in resource-limited settings where healthcare infrastructure is fragile. Continuous education and training of healthcare and veterinary professionals are required, which can be difficult to sustain due to resource limitations. is weak. |
| Enforcing regulations on the prescription, use, and sale of antibiotics in veterinary medicine. This includes promoting antimicrobial stewardship programs and monitoring the usage patterns of antibiotics. | Regulatory enforcement is difficult in regions with weak governance and widespread informal markets, leading to uncontrolled access to antibiotics. Lack of veterinary access coupled with regulation on antibiotic usage would cause unintended misuse or undertreatment causing national and individual economic loss. Low awareness about the dangers of antibiotic misuse among the general population and professionals can undermine regulatory efforts. |
| Promoting a One Health approach to address AMR by fostering collaboration among various sectors, including health, agriculture, environment, and academia. This includes forming national committees, holding regular consultations, and coordinating research efforts. | Aligning the interests and activities of diverse stakeholders across sectors is complex and requires effective communication and leadership. Collaboration amongst sectors would not be fruitful if enough economic assets are not assigned to each sector. |
| Conducting nationwide public awareness campaigns to educate the general population about the dangers of AMR and the importance of responsible antibiotic use. This includes using mass media, social media, and community outreach programs. | Requires economic investments. Also, the lack of veterinary access and economic disparity would counter all the awareness campaigns. Ensuring that public awareness campaigns have a lasting impact requires sustained effort and resources, which can be challenging to maintain over time. Reaching remote and underserved populations can be challenging due to limited access to communication infrastructure and cultural or linguistic barriers. |
| Promoting research on new antibiotics, alternative treatments, and diagnostic tools, as well as operational research on AMR. This includes providing funding, creating research networks, and fostering partnerships between academia and industry. | Research on AMR is expensive and often underfunded, particularly in low-resource settings where competing priorities may limit available funds. |
| Building and upgrading laboratory infrastructure to enhance diagnostic capabilities for detecting AMR. This includes training laboratory personnel, ensuring the availability of necessary equipment, and standardizing diagnostic protocols. | Establishing and maintaining well-equipped laboratories is costly, particularly in low-resource settings. There is often a shortage of trained laboratory personnel, and providing continuous training is resource intensive. |
| Developing and implementing national policies and legislation to combat AMR. This includes drafting new laws, updating existing regulations, and creating national action plans. | Translating legislation into effective action on the ground requires strong institutions and enforcement mechanisms, which may be lacking. |
| Developing systems for the collection, analysis, and sharing of data on antibiotic usage and AMR. This includes creating national databases, ensuring data interoperability, and promoting transparency. | Ensuring that data collected is accurate, consistent, and comprehensive is difficult, particularly in regions with weak data infrastructure. |

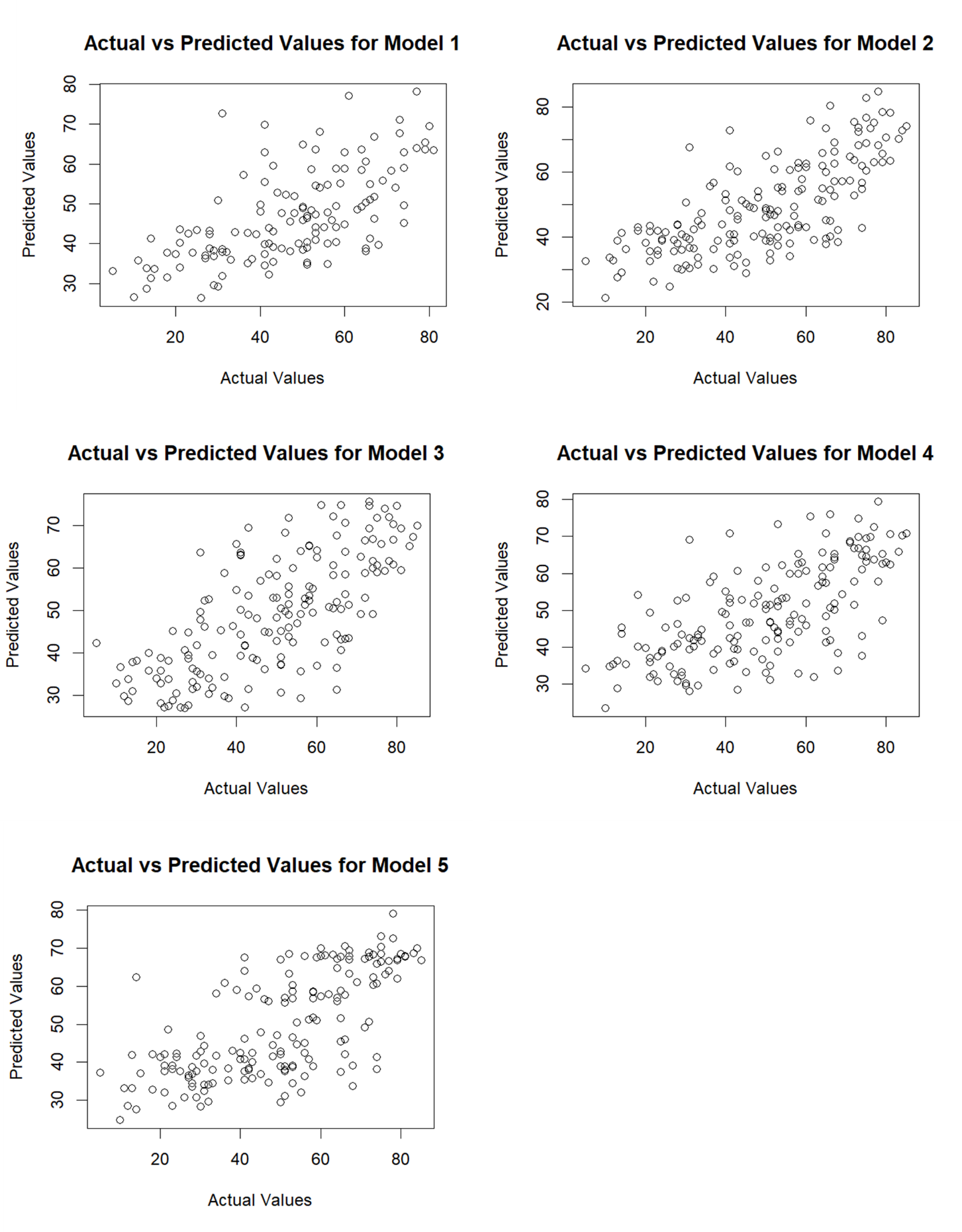

**Figure S1:** This figure illustrates the relationship between actual values (x-axis) and predicted values (y-axis) for Models 1 to 5. Each scatterplot represents one model's performance, highlighting the distribution of data points relative to the ideal prediction line (diagonal). Points closer to this line indicate better agreement between the model's predictions and observed values, whereas deviations reflect prediction errors.

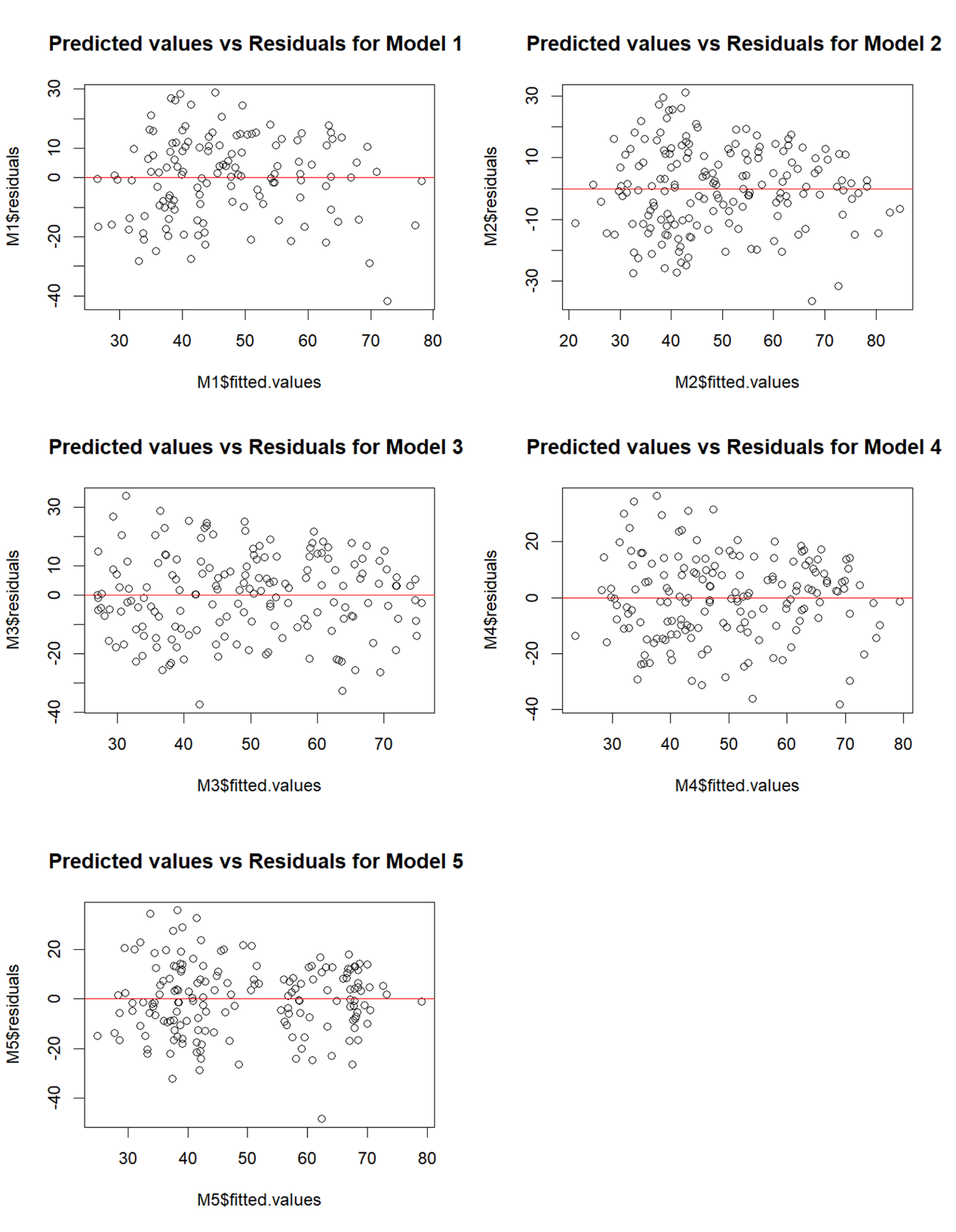

**Figure S2:** Residual plots for five models showing predicted values versus residuals. Each plot corresponds to a specific model (Model 1 to Model 5) and displays the spread of residuals around the horizontal line (y = 0), which indicates the ideal condition of no pattern in residuals. The red lines represent smoothed trend lines for residuals to visualize deviations from randomness. These plots help assess model assumptions, particularly homoscedasticity and independence of residuals.
